# B-type Natriuretic Peptide Changes and Left Ventricular Remodeling Dynamics in Heart Failure with Reduced Ejection Fraction

**DOI:** 10.1101/2025.10.02.25337215

**Authors:** Erick Romero, Yevhen Kushnir, Daniel Mendoza-Quispe, Areej Shahzad, Dev Jaydeep Patel, Kafayat Omadevuae, Padmini Sirish, Nipavan Chiamvimonvat, David A. Liem, Martin Cadeiras

**Author notes:** Address for Correspondence: Martin Cadeiras, MD, Department of Medicine, Division of Cardiovascular Medicine, UC Davis Medical Center, 4860 Y Street, Suite 2820, Sacramento, CA.; or Erick Romero, Department of Internal Medicine, St. Barnabas Hospital, 4422 Third Ave, Bronx, NY, 10457.

## Abstract

**Background:** B-type natriuretic peptide (BNP) is an important biomarker in heart failure with reduced ejection fraction (HFrEF). We aimed to explore changes in BNP and their relationship with long-term dynamics of left ventricular (LV) geometry.

**Methods:** This was a single-center retrospective cohort. Inclusion criteria included LV ejection fraction (LVEF) <40% measured by echocardiography, BNP ≥100 pg/mL at baseline, and a subsequent BNP measure within a year. Percent BNP change from baseline was computed and divided into tertiles. Tertiles represented decreasing, minimal changes, and rising BNP levels. The study endpoint included LV internal dimension at end-systole (LVIDs), LV internal dimension at end-diastole (LVIDd), and LVEF. The secondary endpoint consisted of all-cause mortality.

**Results:** A total of 887 patients were included. Baseline characteristics, including age, sex, blood pressure, atrial fibrillation, baseline BNP, and LVEF, varied among tertiles (p<0.05). When comparing to the rising BNP tertile, the decreasing BNP tertile showed decreased trends of LVIDs (p=0.001), LVIDd (p=0.006); and increased trends of LVEF (p=0.008). All-cause mortality was higher in the rising BNP tertile (p<0.05) compared to the decreasing tertile.

**Conclusion:** In a real-world routine HFrEF cohort, this study demonstrates the *time-dependent* relationship between BNP changes, LV remodeling dynamics, and survival outcomes. Findings contribute to the literature supporting BNP as a dynamic marker for LV remodeling.

**Clinical Perspective:** *What is new?:*

- In a real-world HFrEF cohort, this study demonstrates the *time-dependent* relationship between BNP changes, LV remodeling dynamics, and survival outcomes.

*What are the clinical implications?:*

- Results contribute to the literature supporting early BNP changes as a potential surrogate marker for adverse LV remodeling in a time-dependent fashion.

## INTRODUCTION

B-type natriuretic peptide (BNP) is an important clinical biomarker in heart failure (HF).^1^ Elevated BNP levels are strongly indicative of HF, and are commonly utilized in clinical practice to guide management.^2^ The ventricular myocardium secretes this BNP in response to increased wall stress and pressure overload.^3^ It is a counter-regulatory hormone that promotes vasodilation, natriuresis, and diuresis.^4^

A key pathophysiological process in HF with reduced ejection fraction (HFrEF) is left ventricular (LV) remodeling.^5^ The LV remodeling is marked by progressive dilation, wall thinning, and reduced contractile function.^6^ Importantly, LV remodeling correlates with adverse clinical outcomes.^7,8^ Thus, LV remodeling is an important marker in HFrEF.

While the relationship between natriuretic peptides and LV remodeling exists, the relationship between changes in BNP and time-dependent LV remodeling has not been extensively studied in real-world settings.^6,9,10^ Therefore, the objective of this study was to evaluate the association between changes in BNP and LV remodeling over time in real-world clinical data. We hypothesized that patients who experience greater reductions in BNP levels would demonstrate more favorable LV remodeling and survival outcomes.

## METHODS

### Data source, study design, and inclusion criteria

We conducted a single-center retrospective cohort study using data collected from the University of California, Davis Medical Center. Data was collected through the institution’s electronic health record data warehouse using a time frame between January 2014 to December 2022. Inclusion criteria for the HFrEF cohort consisted of adults aged ≥18 years, International Classification of Diseases (ICD) HF codes (ICD-9 or ICD-10), LVEF ≤40%, and BNP ≥100 pg/mL. Exclusion criteria included patients with ICD codes for cardiac transplant or left ventricular assist device implantation, those without follow-up LVEF values, and those with less than a one-year interval between their first and second BNP measurements. The cohort has been previously validated with a specificity of 0.96 and a sensitivity of 0.60 as compared with physician chart review.^11^

### Baseline definitions

The cohort index date was defined as the date of the first recorded LVEF ≤40%. Demographic variables included age, sex, race, and ethnicity. Baseline laboratory, echocardiogram, and electrocardiogram characteristics were defined as the first value within 90 days or the first available after the indexation date. Comorbidities were defined as diagnoses prior to the cohort index date, and validated comorbidity identification methodologies were used. For this study, we defined guideline-directed medical therapy (GDMT) utilization as the prescription of medications within 6 months prior to and within 3 months after the cohort indexation date. Medications included renin-angiotensin system inhibitors (RASi), β-blockers, mineralocorticoid receptor antagonists (MRA) and sodium-glucose co-transporter 2 inhibitors (SGLT-2). The RASi category comprised angiotensin-converting enzyme inhibitor, angiotensin receptor blocker, and angiotensin receptor neprilysin inhibitor. Patients were placed into one of three dosing categories based on their recorded prescription: either none, <50%, or ≥50% of GDMT target dosing.

### Longitudinal data and cohort stratification

Longitudinal laboratory, electrocardiogram, and echocardiogram variables were extracted, starting from the cohort indexation date until the last available LVEF measurement. After log transformation, percent BNP changes from baseline at one-year were computed, then the cohort was stratified into low, middle, and high BNP change tertiles.

### Study outcomes

The co-primary outcomes were LV internal dimension at end-systole (LVIDs), LV internal dimension at end-diastole (LVIDd), and LVEF, all measured by echocardiography. The secondary outcome included all-cause mortality. All-cause mortality data was retrieved from the institution’s clinical data warehouse. However, cause-specific mortality was not available.

### Statistical analysis

Descriptive statistics were used to summarize baseline characteristics. Between-group comparisons were performed using Wilcoxon signed-rank test for continuous variables or ANOVA, and chi-squared or Fisher exact test for categorical variables as appropriate.

For outcome analysis, linear mixed models were performed to assess changes between BNP change groups and the endpoints over time. A Cox regression model was constructed for survival analysis adjusting for relevant covariates. Analyses were performed using SAS software version 9.4 (SAS institute, Cary, NC, USA).

### Ethics

The university’s Institutional Review Board approved the study, and all data used were de-identified before analysis.

## RESULTS

### Cohort identification and BNP change tertile stratification

An initial 2,334 patients were included. However, in the final cohort, only 887 cases met the inclusion criteria (**Figure 1**). Percent changes in BNP at one-year were categorized into tertiles: low tertile (n=295), middle tertile (n=296), and high tertile (n=296).

**Figure 1.**
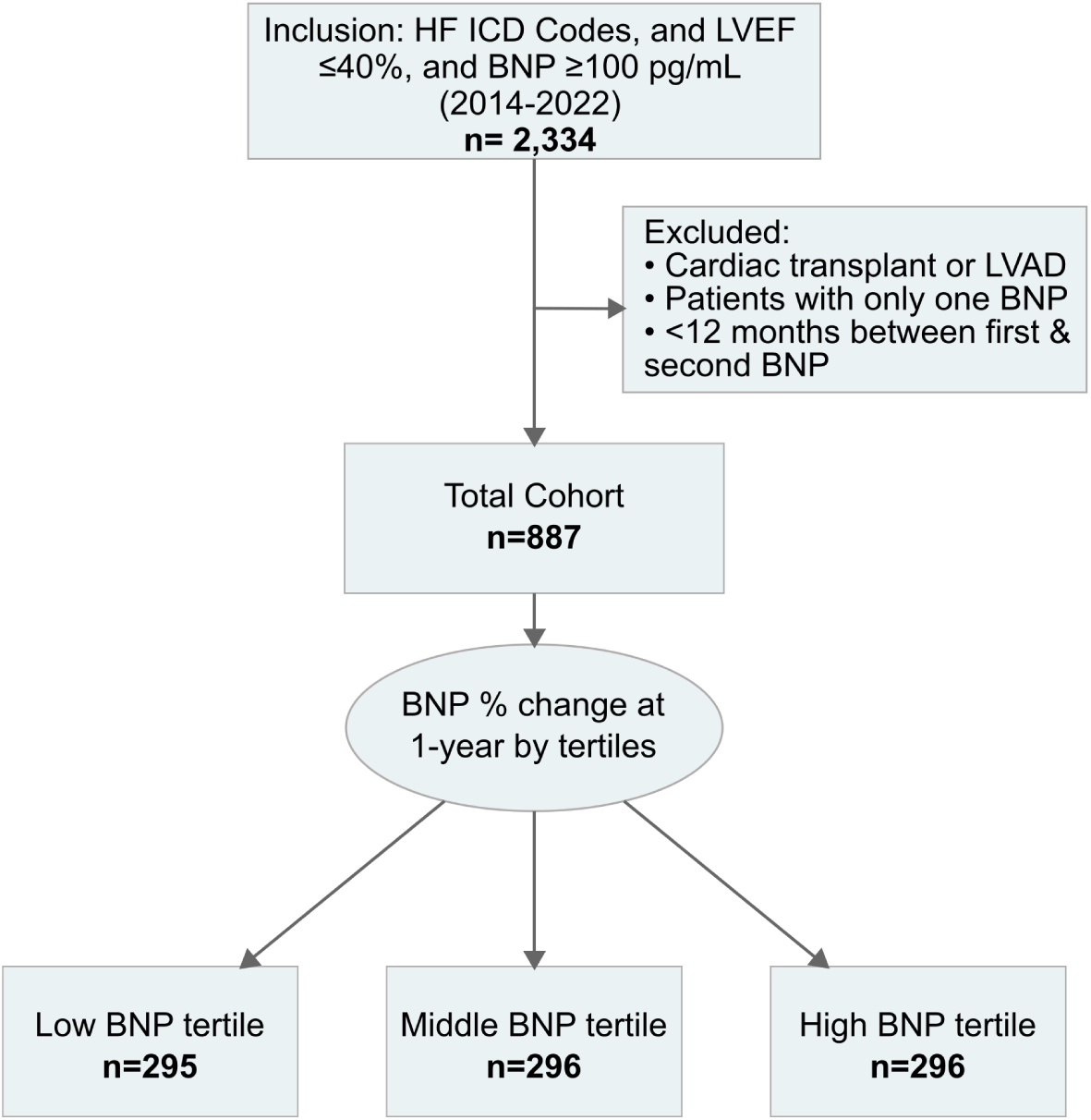
Cohort identification workflow. Abbreviations: BNP = B-type natriuretic peptide; HF = Heart Failure; ICD Codes = International Classification of Diseases codes; LVAD = Left ventricular assist device; LVEF = Left ventricular ejection fraction.

Corresponding to decreasing, minimal changes, and worsening BNP levels (**Figure 1**). The decreasing BNP tertile had a mean BNP reduction of –24% (95% CI, –25% to – 22%); the rising BNP group had a mean increase of 18% (95% CI, 16% – 21%); the minimal BNP change tertile showed a decrease of –4% (95% CI, –4% to – 3%), see **Figure 2A**.

**Figure 2.**
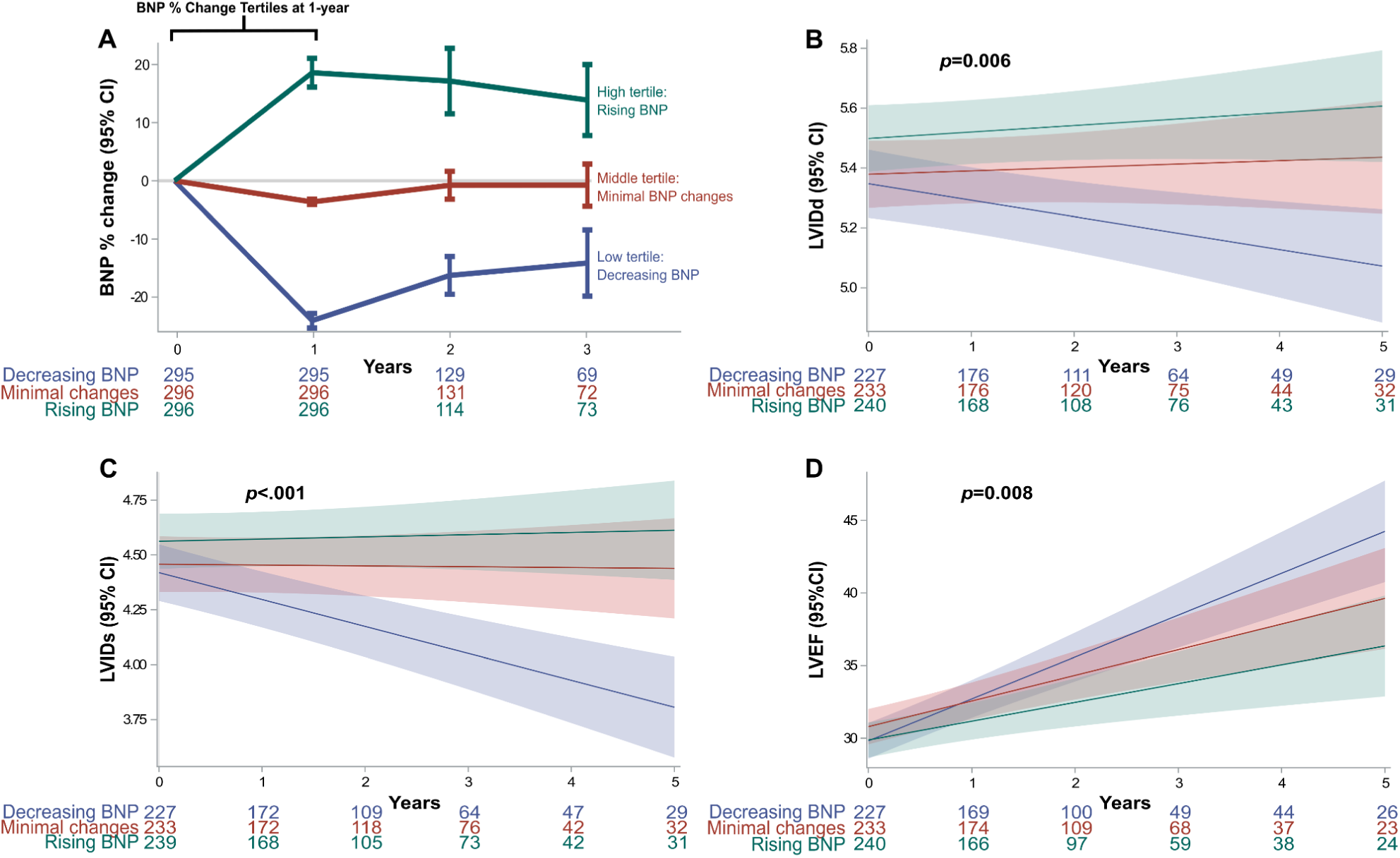
BNP tertile changes and co-primary endpoints. (**A**) Absolute BNP percent change at one-year from baseline: The first year was used to categorize groups based on BNP percent change tertiles. (**B-D**) Co-primary outcomes assessing echocardiographic LV geometry and function showed long-term changes between the BNP change groups. Mixed linear model showed significant differences in LVIDs (p<.006), LVIDs (p<.001) and LVEF (p<.008). Abbreviations: BNP = Brain natriuretic peptide; LV = Left ventricular; LVEF = Left ventricular ejection fraction; LVIDd = LV internal dimension at end-diastole; LVIDs = LV internal dimension at end-systole.

### Baseline characteristics

Baseline characteristics varied across BNP tertiles (**Table 1**). Patients in the decreasing BNP tertile were younger, a median age of 61 years (IQR 53.0-70.5), vs. a median of 65 years (IQR 56.0-77.0) in the rising BNP tertile (p<0.001). Overall, the cohort was predominantly male (68.8%) with a higher male proportion in the minimal changes and rising tertiles groups (73.0% and 69.6%, respectively). Hispanic comprised a 21.4% of the decreasing BNP tertile (p<0.001). Blood pressure and heart rate were elevated in the decreasing BNP tertile group (80.0 mmHg, p=0.022; and 91.0 bpm, p=0.004, respectively). The minimal BNP changes group presented the highest rates of atrial fibrillation patients (41.2%, p=0.007).

**Table 1.** Patient baseline characteristics at the time of first LVEF ≤40% and BNP≥100 pg/mL.

| Median (IQR) or No. (%) |  |  |  |  |  |  |
| --- | --- | --- | --- | --- | --- | --- |
| Characteristics | Miss | Overall (n=887) | Low tertile,<br>decreasing BNP<br>(n=295) | Middle tertile,<br>minimal BNP<br>changes (n=296) | High tertile,<br>rising BNP<br>(n=296) | P* |
| Age, years | 0 | 64 (55-75) | 61 (53.0-70.5) | 65 (57-77) | 65 (56-77) | <.001 |
| **Sex** | 0 |  |  |  |  | 0.049 |
| Male |  | 610 (68.8) | 188 (63.7) | 216 (73.0) | 206 (69.6) |  |
| **Race** | 0 |  |  |  |  | 0.075 |
| White |  | 506 (57.0) | 148 (50.2) | 187 (63.2) | 171 (57.8) |  |
| Black |  | 159 (17.9) | 61 (20.7) | 46 (15.5) | 52 (17.6) |  |
| Asian |  | 46 (5.2) | 13 (4.4) | 14 (4.7) | 19 (6.4) |  |
| Hawaiian/Native American |  | 23 (2.6) | 7 (2.4) | 7 (2.4) | 9 (3.0) |  |
| Other |  | 146 (16.4) | 63 (21.3) | 40 (13.5) | 43 (14.5) |  |
| Unavailable |  | 7 (0.8) | 3 (1.0) | 2 (0.7) | 2 (0.7) |  |
| **Ethnicity** | 0 |  |  |  |  | <.001 |
| Non-Hispanic |  | 761 (85.8) | 229 (77.6) | 271 (91.6) | 261 (88.2) |  |
| Hispanic |  | 120 (13.5) | 63 (21.4) | 24 (8.1) | 33 (11.1) |  |
| Unavailable |  | 6 (0.7) | 3 (1.0) | 1 (0.3) | 2 (0.7) |  |
| **Vitals/Biometrics** |  |  |  |  |  |  |
| Systolic BP, mmHg | 3 | 125 (111-142) | 126 (110-143) | 121 (109-139) | 128 (113-143.8) | 0.100 |
| Diastolic BP, mmHg | 3 | 77 (67-89) | 80 (68-92) | 76 (66-87) | 76 (68-88) | 0.022 |
| MAP, mmHg | 3 | 94 (82.7-105.3) | 96.7 (82.2-106.3) | 90.7 (81.8-103.7) | 94 (84.1-105.0) | 0.068 |
| Heart Rate, beats/min | 1 | 89 (77-102) | 91 (80-105) | 88 (76-102) | 86 (75-99.2) | 0.004 |
| BMI, kg/m<sup>2</sup> | 91 | 28.4 (24.6-33.4) | 28.2 (24.9-32.9) | 28.7 (24.8-32.8) | 28.2 (24.3-33.8) | 0.934 |
| **Medical History** |  |  |  |  |  |  |
| Hypertension | 8 | 639 (72.7) | 215 (73.4) | 201 (69.1) | 223 (75.6) | 0.198 |
| Diabetes | 8 | 402 (45.7) | 130 (44.4) | 123 (42.3) | 149 (50.5) | 0.114 |
| Hyperlipidemia | 8 | 390 (44.4) | 123 (42.0) | 125 (43.0) | 142 (48.1) | 0.271 |
| Ischemic Heart Disease | 8 | 325 (37.0) | 96 (32.8) | 118 (40.5) | 111 (37.6) | 0.144 |
| Atrial Fibrillation | 8 | 301 (34.2) | 86 (29.4) | 120 (41.2) | 95 (32.2) | 0.007 |
| Chronic Kidney Disease | 8 | 244 (27.8) | 77 (26.3) | 77 (26.5) | 90 (30.5) | 0.432 |
| **Laboratory** |  |  |  |  |  |  |
| BNP, pg/mL | 0 | 663 (298-1305) | 837 (419-1553) | 698<br>(362.5-1344.5) | 433.5<br>(139.2-981.0) | <.001 |
| NT-proBNP, pg/mL | 777 | 1259.5<br>(353.8-2896.2) | 922 (237-2520) | 2045.5<br>(952.5-3485.0) | 874<br>(355.5-2414.5) | 0.056 |
| Sodium, mEq/L | 0 | 137 (135-139) | 137 (135-139) | 137 (135-139) | 137.5 (135-139) | 0.675 |
| Potassium, mEq/L | 0 | 4 (3.7-4.4) | 3.9 (3.6-4.2) | 4.1 (3.7-4.4) | 4 (3.7-4.4) | 0.003 |
| Creatinine, mg/dl | 0 | 1.2 (0.9-1.6) | 1.1 (0.9-1.5) | 1.2 (1.0-1.6) | 1.1 (0.9-1.5) | 0.101 |
| eGFR, ml/min/1.73 m<sup>2</sup> | 39 | 56 (45-60) | 57 (48-67) | 56 (43.5-60) | 55 (44-60) | 0.182 |
| **Echocardiogram** |  |  |  |  |  |  |
| LVEF, % | 0 | 30 (25-35) | 30 (20-35) | 30 (25-40) | 30 (25-35) | <.001 |
| IVSd, cm | 1 | 1.2 (1.0-1.3) | 1.1 (1.0-1.3) | 1.2 (1.0-1.3) | 1.2 (1.0-1.3) | 0.140 |
| LVIDd, cm | 2 | 5.7 (5.1-6.3) | 5.7 (5.1-6.3) | 5.6 (5.1-6.3) | 5.6 (5.1-6.2) | 0.376 |
| LVIDs, cm | 6 | 4.8 (4.1-5.5) | 4.9 (4.2-5.6) | 4.7 (4.1-5.5) | 4.7 (4.1-5.3) | 0.203 |
| PASP, mmHg | 56 | 41.8 (31.7-50.2) | 42.6 (31.8-50.1) | 42.5 (33.0-51.2) | 38.9 (29.7-49.4) | 0.113 |
| PW, cm | 2 | 1.1 (1.0-1.3) | 1.1 (1.0-1.3) | 1.1 (1.0-1.3) | 1.1 (1.0-1.3) | 0.195 |
| TAPSE, cm | 19 | 1.7 (1.4-2.1) | 1.7 (1.3-2.1) | 1.7 (1.4-2.0) | 1.8 (1.5-2.1) | 0.046 |
| **Electrocardiogram** |  |  |  |  |  |  |
| QTc, ms | 8 | 500 (474-531) | 497 (474.8-530.2) | 503 (476.0-534.8) | 499 (471-528) | 0.203 |
| **GDMT & target dose<sup>Δ</sup>** |  |  |  |  |  |  |
| RASi<sup>§</sup> | 147 |  |  |  |  | 0.022 |
| none |  | 139 (18.8) | 47 (18.6) | 57 (22.5) | 35 (15.0) |  |
| <50% target |  | 309 (41.8) | 97 (38.3) | 116 (45.8) | 96 (41.0) |  |
| ≥50% target |  | 292 (39.5) | 109 (43.1) | 80 (31.6) | 103 (44.0) |  |
| β-Blocker | 147 |  |  |  |  | 0.061 |
| none |  | 270 (36.5) | 85 (33.6) | 83 (32.8) | 102 (43.6) |  |
| <50% target |  | 247 (33.4) | 82 (32.4) | 93 (36.8) | 72 (30.8) |  |
| ≥50% target |  | 223 (30.1) | 86 (34.0) | 77 (30.4) | 60 (25.6) |  |
| MRA | 147 |  |  |  |  | 0.057 |
| none |  | 455 (61.5) | 143 (56.5) | 155 (61.3) | 157 (67.1) |  |
| <50% target |  | 0 | 0 | 0 |  |  |
| ≥50% target |  | 285 (38.5) | 110 (43.5) | 98 (38.7) | 77 (32.9) |  |
| SGLT2i | 147 | 62 (8.4) | 26 (10.3) | 21 (8.3) | 15 (6.4) | 0.306 |
| GDMT Triple therapy | 147 | 149 (20.1) | 60 (23.7) | 50 (19.8) | 39 (16.7) | 0.150 |
| GDMT Triple therapy, ≥50% target | 147 | 40 (5.4) | 21 (8.3) | 9 (3.6) | 10 (4.3) | 0.040 |
| Loop diuretics | 0 | 866 (97.6) | 290 (98.3) | 290 (98.0) | 286 (96.6) | 0.362 |

\* p-values comparing all BNP tertile groups using Chi-squared or Kruskal-Wallis
<sup>Δ</sup> Target doses of guideline directed medical therapy (GDMT) as recommended in treatment guidelines.
<sup>§</sup> RASi consisted of angiotensin-converting enzyme inhibitor, angiotensin receptor blocker, angiotensin receptor neprilysin inhibitor.
**Abbreviations:** BMI = Body Mass Index; BNP = B-type natriuretic peptide; BP = Blood Pressure; eGFR = Estimated glomerular filtration rate; GDMT = Guideline-directed medical therapy; IVSd= Interventricular septum thickness at end-diastole; LVEDd = Left ventricle end diastolic diameter; LVEF = Left ventricular ejection fraction; LVIDd = Left ventricular internal dimension at end -diastole; LVIDs = Left internal dimension at end -systole; MAP = Mean arterial pressure; MRA = Mineralocorticoid receptor antagonist; NT-proBNP = N-terminal (NT)-pro hormone BNP; PASP = Pulmonary artery systolic pressure; PW = Left ventricular posterior wall; QTc = QT corrected for heart rate; RASi = Renin-angiotensin-system inhibitor; SGLT2i = Sodium glucose co-transporter 2 inhibitors; TAPSE = Tricuspid annular plane systolic excursion.

The decreasing BNP group had elevated BNP levels, a median of 837.0 pg/mL (IQR 419.0-1553.0) when compared to the other tertiles (p< 0.001). Potassium levels were elevated in the minimal changes and rising BNP tertiles (p=0.003). Minimal changes and rising BNP tertiles had a higher LVEF. Baseline TAPSE was also higher in the rising BNP tertile (p=0.046).

The use of GDMT analysis showed that RASi were prescribed more frequently in the rising BNP group. However, the decreasing BNP group had a higher use of GDMT triple therapy at ≥50% target doses (8.3%, p=0.040).

### Primary outcomes: changes in LV geometry and function

Linear mixed models for LVIDd different trajectories between tertiles (p=0.006). The decreasing BNP group demonstrated a steady decrease in LVIDd, **Figure 2B**. Similarly, LVIDs steadily decreased in the decreasing BNP tertile (p<0.001), **Figure 2C**. Finally, the decreasing BNP tertile exhibited an improvement in LVEF over time (p=0.008), **Figure 2D**.

### Secondary outcome: all-cause mortality

Cox regression analysis adjusted for clinically relevant covariates resulted in worse survival outcomes for the rising BNP tertile when compared to decreasing BNP (p=0.009) and minimal BNP changes (p=0.02), see **Figure 3**. Clinically relevant covariates, includined age, years, sex, BMI, heart rate, atrial fibrillation, baseline BNP, creatinine, and LVEF.

**Figure 3.**
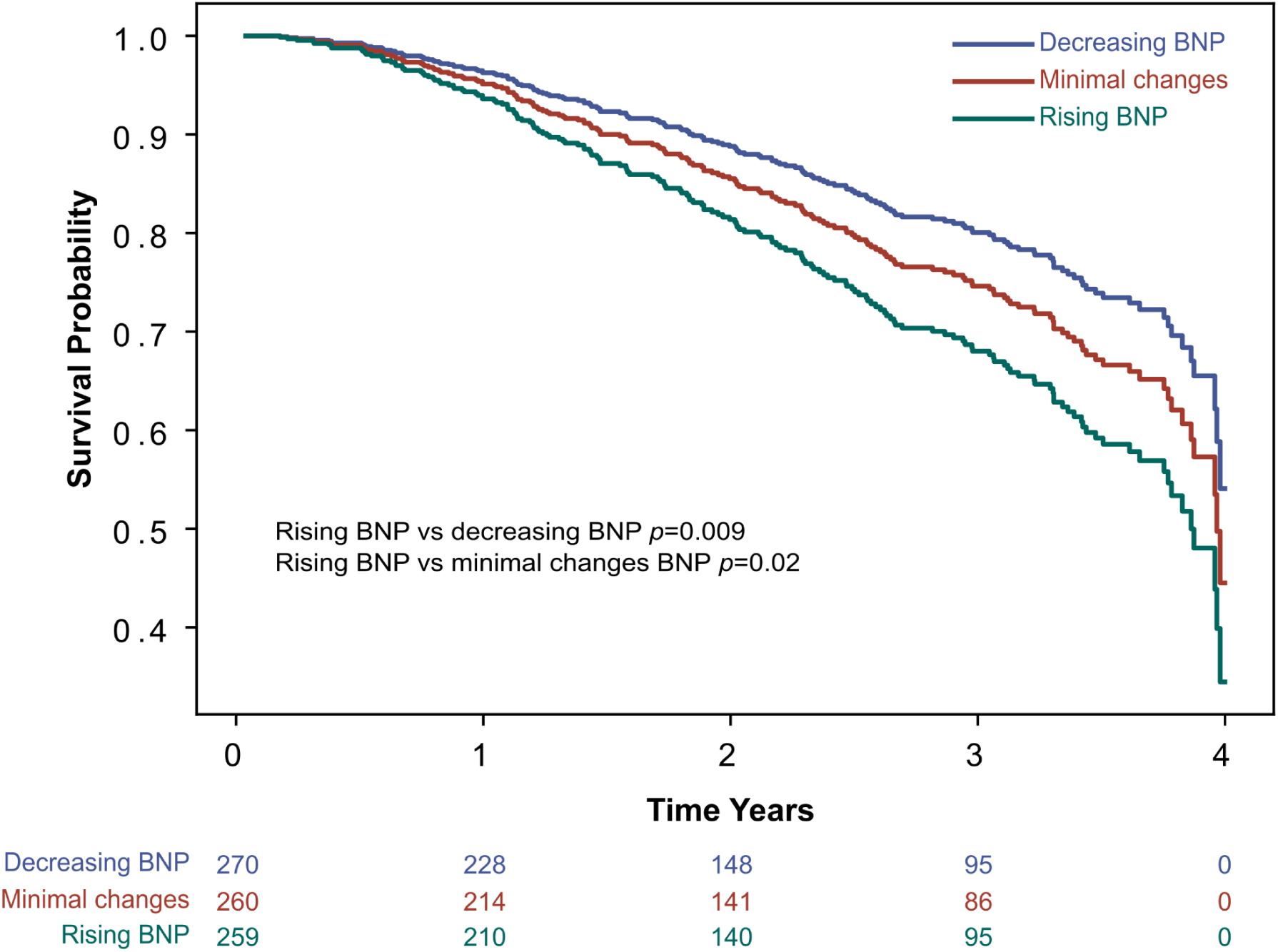
Secondary endpoint: all-cause mortality. Survival curve for all-cause mortality by BNP tertile change groups, adjusted for covariates in a Cox regression model. The Cox regression model was adjusted for clinically relevant covariates, including age, years, sex, BMI, heart rate, atrial fibrillation, baseline BNP, creatinine, and LVEF. Abbreviations: BMI = Body mass index; BNP = Brain natriuretic peptide; LVEF = Left ventricular ejection fraction.

## DISCUSSION

In this retrospective cohort, the results suggest an association between one-year changes in BNP levels and long-term LV remodeling in patients with HFrEF. Specifically, patients who experienced early reductions in BNP levels demonstrated favorable long-term LV remodeling and improved overall mortality outcomes. Findings underscore BNP changes as a dynamic marker for LV function.

### Cohort characteristics and BNP tertile interpretation

Baseline characteristics in **Table 1**, provide insights into the varying profiles across BNP tertiles. Compared to other groups, the decreasing BNP group comprised younger, higher proportion of females, and Hispanics, and higher blood pressure and heart rate values. In addition, it presented the lowest atrial fibrillation rates. In terms of HF severity, the decreasing BNP tertile had higher BNP levels at baseline, worse LVEF, and lower TAPSE compared to other groups. Finally, the use of RASi and GDMT Triple therapy at ≥50% was higher in this group. Taken together, the decreasing BNP group is indicative of younger patients with more severe HF but a better compensatory state and/or greater responsiveness to treatment.^12^

### Interpretation of primary and secondary endpoints

The outcome analysis supports the hypothesis that BNP reductions at one-year is associated with favorable long-term LV remodeling and overall survival. The decreasing BNP tertile, characterized by substantial reductions in BNP from the indexation cohort date, showed significantly better LVIDd, LVIDs, and LVEF trajectories. This suggests that BNP changes may serve as an early predictive marker for long-term LV function. These findings are consistent with prior studies in myocardial infarction cohorts, where BNP has been shown to predict LV remodeling over time.^13,6^

BNP is a cardiac natriuretic peptide hormone primarily produced by ventricular myocytes. The primary stimulus for synthesis and secretion is myocyte stretch. The biological effects include diuresis, natriuresis, vasodilatation, inhibition of renin and aldosterone production, and regulation of cardiac and vascular myocyte growth.^14^ BNP acts as an endogenous brake on signaling pathways that drive the progression from LV hypertrophy through remodeling, heart failure, and death.^15^ Multiple studies have demonstrated BNP’s anti-hypertrophic effects in both exogenous and endogenous experimental settings.^16,17^

Prior studies have shown that BNP levels can be valuable for prognosis. One study found that in a cohort of MI survivors over six months, patients with LV remodeling had higher levels of BNP on days 7, 90, and 180 compared to those without LV remodeling.^18^ Another study reported that in a cohort of MI survivors over one year, N-BNP levels were higher in patients with larger myocardial scars, which are identified as a major determinant of LV remodeling.^19^

Due to the mechanisms triggering BNP production and its biological effects, elevated BNP levels can reflect both LV systolic dysfunction and the body’s compensatory response.^20^ These findings suggest that a BNP downtrend may be associated with improved systolic function, leading to a decrease in LV end-diastolic volume, reduced LV wall stress, and diminished BNP production. This can explain why patients with greater BNP reductions over the course of a year exhibited decreased LVIDs, lower LVIDd, and increased LVEF, which can be surrogate signs of LV remodeling improvement. Improved BNP and LV function directly translates into better survival outcomes.

### Clinical implications

In this study, we demonstrated that patients in the low BNP tertile had reduced LVIDs and LVIDd and increased LVEF compared to those in the high BNP tertile. These findings suggest that BNP dynamic reduction is indicative of LV remodeling improvement in a time-dependent fashion. Therefore, contributing to the literature of the relationship of BNP and LV remodeling dynamics.

### Study limitations

Limitations should be considered, the findings may not be generalizable to other populations due the single-center retrospective design. In addition, findings should be interpreted as associations rather than causal relationships. Cohort identification required follow-up data, which may select more compliant or healthier patients. Cause-specific mortality data was not collected in our institution, cause specific mortality analysis was not feasible.

## CONCLUSION

In a real-world routine HFrEF cohort, his study suggests that BNP changes within one year are associated with LV remodeling dynamics in the long term. Greater BNP reductions were associated with more favorable LV function, structural remodeling, and better all-cause mortality. Findings contribute to the growing literature of BNP as a dynamic marker for LV remodeling.

## Data Availability

Data will be available upon request.

## Abbreviations

BNP: B-type natriuretic peptide
GDMT: Guideline-directed medical therapy
HF: Heart failure
HFrEF: Heart failure with reduced ejection fraction
ICD: International Classification of Diseases
LV: Left ventricular
LVAD: Left ventricular assist device
LVEF: Left ventricular ejection fraction
LVIDd: Left ventricular internal dimension at end-diastole
LVIDs: Left ventricular internal dimension at end-systole
MRA: Mineralocorticoid receptor antagonists
NT-proBNP: N-terminal pro-B-type natriuretic peptide
RASi: Renin-angiotensin system inhibitors
SGLT-2: Sodium-glucose co-transporter 2 inhibitors

## REFERENCES

1. Chow SL, Maisel AS, Anand I, et al. Role of Biomarkers for the Prevention, Assessment, and Management of Heart Failure: A Scientific Statement From the American Heart Association. Circulation. 2017;135(22):e1054–e1091. doi:10.1161/CIR.0000000000000490

2. Heidenreich PA, Bozkurt B, Aguilar D, et al. 2022 AHA/ACC/HFSA Guideline for the Management of Heart Failure: A Report of the American College of Cardiology/American Heart Association Joint Committee on Clinical Practice Guidelines. Journal of the American College of Cardiology. 2022;79(17):e263–e421. doi:10.1016/j.jacc.2021.12.012

3. Ibrahim NE, Januzzi JL. Established and Emerging Roles of Biomarkers in Heart Failure. Circulation Research. 2018;123(5):614–629. doi:10.1161/CIRCRESAHA.118.312706

4. Kuwahara K. The natriuretic peptide system in heart failure: Diagnostic and therapeutic implications. Pharmacology & Therapeutics. 2021;227:107863. doi:10.1016/j.pharmthera.2021.107863

5. Frantz S, Hundertmark MJ, Schulz-Menger J, Bengel FM, Bauersachs J. Left ventricular remodelling post-myocardial infarction: pathophysiology, imaging, and novel therapies. European Heart Journal. 2022;43(27):2549–2561. doi:10.1093/eurheartj/ehac223

6. Aimo A, Gaggin HK, Barison A, Emdin M, Januzzi JL. Imaging, Biomarker, and Clinical Predictors of Cardiac Remodeling in Heart Failure With Reduced Ejection Fraction. JACC: Heart Failure. 2019;7(9):782–794. doi:10.1016/j.jchf.2019.06.004

7. Xu L, Pagano J, Chow K, et al. Cardiac remodelling predicts outcome in patients with chronic heart failure. ESC Heart Failure. 2021;8(6):5352. doi:10.1002/ehf2.13626

8. Lindman BR, Asch FM, Grayburn PA, et al. Ventricular Remodeling and Outcomes After Mitral Transcatheter Edge-to-Edge Repair in Heart Failure. JACC: Cardiovascular Interventions. 2023;16(10):1160–1172. doi:10.1016/j.jcin.2023.02.031

9. Daubert MA, Adams K, Yow E, et al. NT-proBNP Goal Achievement Is Associated With Significant Reverse Remodeling and Improved Clinical Outcomes in HFrEF. JACC: Heart Failure. 2019;7(2):158–168. doi:10.1016/j.jchf.2018.10.014

10. Shiba M, Kato T, Morimoto T, et al. Changes in BNP levels from discharge to 6-month visit predict subsequent outcomes in patients with acute heart failure. PLoS ONE. 2022;17(1):e0263165. doi:10.1371/journal.pone.0263165

11. Romero E, Baltodano AF, Rocha P, et al. Clinical, Echocardiographic, and Longitudinal Characteristics Associated With Heart Failure With Improved Ejection Fraction. The American Journal of Cardiology. 2024;211:143–152. doi:10.1016/j.amjcard.2023.10.086

12. Regan JA, Kitzman DW, Leifer ES, et al. Impact of Age on Comorbidities and Outcomes in Heart Failure With Reduced Ejection Fraction. JACC: Heart Failure. 2019;7(12):1056–1065. doi:10.1016/j.jchf.2019.09.004

13. Bauters C, Fertin M, Pinet F. B-type natriuretic peptide for the prediction of left ventricular remodelling. Cardiovasc J Afr. 2014;25(1):33–39.

14. Hall C. Essential biochemistry and physiology of (NT-pro)BNP. Eur J Heart Fail. 2004;6(3):257–260. doi:10.1016/j.ejheart.2003.12.015

15. Ritchie RH, Rosenkranz AC, Kaye DM. B-type natriuretic peptide: endogenous regulator of myocardial structure, biomarker and therapeutic target. Curr Mol Med. 2009;9(7):814–825. doi:10.2174/156652409789105499

16. Sarzani R, Allevi M, Di Pentima C, Schiavi P, Spannella F, Giulietti F. Role of Cardiac Natriuretic Peptides in Heart Structure and Function. Int J Mol Sci. 2022;23(22):14415. doi:10.3390/ijms232214415

17. Sangaralingham SJ, Kuhn M, Cannone V, Chen HH, Burnett JC. Natriuretic peptide pathways in heart failure: further therapeutic possibilities. Cardiovasc Res. 2022;118(18):3416–3433. doi:10.1093/cvr/cvac125

18. Hsu JT, Chung CM, Chu CM, et al. Predictors of Left Ventricle Remodeling: Combined Plasma B-type Natriuretic Peptide Decreasing Ratio and Peak Creatine Kinase-MB. International Journal of Medical Sciences. 2017;14(1):75–85. doi:10.7150/ijms.17145

19. Orn S, Manhenke C, Squire IB, Ng L, Anand I, Dickstein K. Plasma MMP-2, MMP-9 and N-BNP in long-term survivors following complicated myocardial infarction: relation to cardiac magnetic resonance imaging measures of left ventricular structure and function. J Card Fail. 2007;13(10):843–849. doi:10.1016/j.cardfail.2007.07.006

20. Goetze JP, Bruneau BG, Ramos HR, Ogawa T, de Bold MK, de Bold AJ. Cardiac natriuretic peptides. Nat Rev Cardiol. 2020;17(11):698–717. doi:10.1038/s41569-020-0381-0

